# Dual Burden of Malnutrition Among Mother-Child Dyads in Mozambique: Findings from the Demographic and Health Survey 2022-2023

**DOI:** 10.64898/2026.08.07.26359956

**Authors:** Nahin Shakurun, Fernanda Andre, Nazeem Muhajarine

## Abstract

**Introduction:** Nutritional challenges are a global public health concern, especially among children under five in sub-Saharan African countries. The coexistence of an overweight/obese mother and an underweight child in the same household is recognized as a dual burden of malnutrition (DBM). Our study aimed to examine the prevalence and associated factors of DBM among mother-child dyads in Mozambique.

**Methods:** We used nationally representative, cross-sectional data from the Mozambique Demographic and Health Survey 2022-2023 (n=3,605 mother-child dyads). The children’s undernutrition condition and maternal BMI status were calculated using the WHO standard reference guidelines. The outcome variable, dual burden of malnutrition, was then created if the children had any undernutrition condition (stunting, wasting, or undernutrition) and the corresponding mother was overweight/obese. Multivariable binary logistic regression, Erreygers’s concentration index, and concentration curve were analyzed to determine associated factors and social inequalities.

**Results:** The prevalence of the dual burden of malnutrition was about 5.51%. Mothers aged *≥*34 years [aOR (95% CI): 4.01(1.44, 11.14); p<0.05] and mothers with four or more children [aOR (95% CI): 2.68(1.29, 5.57); p<0.05] had higher odds of DBM. Rural residence and using unimproved toilet facilities (latter an indicator) were also significantly associated with experiencing DBM. Additionally, maternal age modified the association between women’s empowerment and mother-child DBM. Women aged 15–19 years at the lowest and highest empowerment levels were more likely to experience DBM compared to women 20 years or older. A positive and statistically significant concentration index indicates that wealth-related inequalities exist, with DBM more concentrated among wealthier mother-child dyads.

**Conclusion:** Our study highlights the persistence of household-level DBM in Mozambique. These findings emphasize the need for targeted interventions addressing social and economic inequalities, including poor sanitation marker of broader household deprivation. Prioritizing integrated maternal-child nutrition interventions within national strategies is essential to improving equity in nutrition and health outcomes.

## INTRODUCTION

Until recently, undernutrition and overnutrition were traditionally considered two distinct public health concerns. However, global evidence now demonstrates that both undernutrition and overnutrition can coexist within the same community, families, or individuals, even.(1,2) As the global nutrition transition accelerates, an increasing proportion of individuals are exposed to multiple forms of malnutrition throughout their lives and directly bear the dual burden of malnutrition (DBM).(3,4) The World Health Organisation (WHO) defines DBM as ‘the coexistence of undernutrition along with overweight, obesity-related or diet-related non-communicable diseases (NCDs), within individuals, households, and populations, and across the life course.’(5) At an individual level, this coexistence emerges as a combination of impaired growth and development, and overweight or obesity in children.(6) Household-level DBM refers to the simultaneous presence of undernourishment in one member and overnutrition in another within the same household. The coexistence of both ends of the malnutrition spectrum within the same community, particularly amongst children, is referred to as the community-level DBM. (1,4)

DBM is a significant public health issue faced by many nations, and it is closely linked to several Sustainable Development Goals (SDGs). One of the most crucial intersections between good nutrition and the SDGs is target number 2.(7) The United Nations (UN) SDGs target 2.0 (SDG-2) aims to eradicate hunger and ensure that all people, particularly those living in poverty and people in other vulnerable conditions, including infants and children under five, have access to sufficient and nutritious food. The goal is to achieve food security and improve nutrition throughout the year.(8) With a specific focus on the African region, the WHO has underscored the importance of addressing DBM to achieve the UN’s nutrition and SDGs from 2019 to 2025.^1,9^

Global estimates reveal the alarming prevalence and impact of malnutrition worldwide. Every year, millions of children under five suffer from impaired growth and development, while millions more succumb to overweight or obesity-related complications.(1,9) These figures demonstrate that one in three children in low- and middle-income countries (LMICs) is affected by DBM. For several decades, maternal and child malnutrition have been a major public health concern in LMICs.(9,10) However, lifestyle habits and health-seeking behavioural changes coupled with increased urbanisation and globalisation have led to a significant nutritional transition pattern from underweight to overweight in these settings.(11) Although once considered uncommon to find the coexistence of undernutrition and overnutrition within the same country, community, and household, these countries continue to face DBM, nutrition-related NCDs, food insecurity, and continuing threats of infectious diseases.(12) In 2020, undernutrition contributed to approximately 45% of deaths amongst children under five. Additionally, it is estimated that stunting affected 149 million children in this age group, while wasting impacted over 49 million children.(9)

In sub-Saharan Africa, 20 to 50% of the urban population is overweight or obese, with global estimates indicating that by 2025, 75% of the world’s obese population may live in low-income countries.(13,14) Further, women of reproductive age (WRA) who experience DBM are at higher risk of experiencing adverse maternal outcomes for themselves and their offspring during preconception, pregnancy, and breastfeeding.(15–17) The prevalence of underweight amongst WRA in Africa has declined considerably over the last few decades, but it remains higher than 10% in the region.(18) However, evidence indicates a rising trend in overweight across LMICs.(18) National and regional studies conducted in Africa revealed that DBM affects 14% of WRA in Ghana. The prevalence of undernutrition is reported to be 4% in Ghana, 11% in Tanzania, 16.9% in Kenya, and 25.5% in Ethiopia.(19–22) Furthermore, about 27% of women in Mali and 63% in Ethiopia are affected by overnutrition (overweight or obesity).(20) Other studies also found that the magnitude of overweight and obesity, respectively, was 28.1% and 7.7% in Kenya, while in Ghana, it was 21% and 12%.(23,24)

A meta-analysis of demographic and health surveys conducted to assess DBM in LMICs, revealed that Mozambique has one of the highest rates of stunting amongst children under five, estimated at 43.0%. Other countries across the sub-Saharan African region with similar high prevalences of stunting in this age group include Burundi (56.0%), Madagascar (50.0%), the Democratic Republic of Congo (43.0%), and Chad (40.0%). Additionally, countries experiencing the highest coexistence of child undernutrition alongside overweight or obesity include South Africa, Sao Tome and Principe, Eswatini, Comoros, and Equatorial Guinea.(25)

Moreover, studies have consistently found that women are more likely to be underweight or overweight compared to men. This discrepancy may stem from factors such as the nature of reproductive health, physical activity levels, lower social status, experiencing poverty, and limited access to formal and quality education. These studies also revealed that maternal undernutrition was associated with factors such as occupation, marital status, contraceptive use, region, residence, and community-level wealth.(2,26,27)

Mozambique, a country in the southeastern sub-Saharan Africa region, grapples with a high burden of malnutrition despite governmental and international efforts to reduce hunger and undernutrition. This highlights the need for ongoing public health interventions at the national level to effectively address this pressing public health issue. Addressing this issue of DBM in Mozambique requires understanding the complex sociodemographic characteristics affecting mother-child dyads. The coexistence of undernutrition and overnutrition within the same families has not been fully explored using nationally representative data. The country’s weak health infrastructure may worsen its population’s nutritional status and outcomes and impede development. Furthermore, to achieve target 2.0 of the SDGs, which focuses on reducing all forms of malnutrition at global, regional, national, and subnational levels, it is critical to understand the sociodemographic drivers of DBM.(28) The current study aimed, therefore, to estimate the prevalence of DBM and assess the socioeconomic inequalities contributing to this significant public health issue amongst mothers and their children under five years old in Mozambique.

## METHODS

### Data source

The Mozambique Demographic and Health Survey (DHS) 2022-23 dataset was used for this study.(29) It was a nationwide cross-sectional survey, encompassing ten provinces (Niassa, Cabo Delgado, Nampula, Zambezia, Tete, Manica, Sofala, Inhambane, Gaza, and Maputo), and the city of Maputo, which holds provincial status as the country’s capital city. Data collection was conducted employing a two-stage stratified sampling design. In the first stage, a total of 619 enumeration areas (EAs) were selected with probability proportional to size, measured by the number of households in each explicit stratum, based on the 2017 IV General Census of Population and Housing (IV RGPH). In the second stage, 26 households were systematically selected with equal probabilities from each EA. Based on this procedure, 16,045 households were selected for data collection. All women aged 15-49 years who were usual residents or visitors in the household the night before the interviews were eligible for the interview. For our analysis, we have used the children recode (MZKR) data file. A total of 9,289 mothers who had at least one child under five years old were included in the study. However, women who were currently pregnant were excluded from the study. Finally, 3,605 mother-child dyads were included in our analysis, whose weights and heights were measured. Details of the participants’ selection for this analysis are presented in **supplementary figure S1**.

### Variables

#### Outcome variable

The outcome variable was dual burden of malnutrition (DBM). In this study, participants were categorized as experiencing DBM if the child was experiencing stunting, wasting or was underweight and the mother was overweight/obese using WHO standards.(30) Although DBM can also occur in the reverse form (i.e., underweight mother and overweight/obese child), this form was not examined due to its low prevalence [Underweight mothers (5.02%, n= 181) and overweight/obese child (0.8%, n= 28)] in the study sample, which limits statistical power for meaningful analysis.

Anthropometric data on height and weight, collected in the survey, were used to measure the nutritional status of children and mothers. Weights were measured using the Seca 878 digital scale.^29^ Heights were measured using an altimeter with ShorrBoard reference. Children under 24 months of age were measured in lying position, while children 24-59 months and mothers were measured in standing position.(29) For DHS data, the WHO 2006 children growth standard was used to calculate a standard deviation (SD) z-score.(31) Stunting, wasting, and underweight conditions for children were defined if height-for-age, weight-for-height, and weight-for-age z-scores below -2 SD of the WHO child growth standards median, respectively.(31) The measurements were considered invalid if height-for-age and weight-for-age z-scores below -6 SD and weight-for-height z-scores below -5 SD.(31) Maternal body mass index (BMI) was calculated as weight in kilograms (kg) divided by height in square meters. Maternal BMI was categorized into three groups: Underweight (BMI<18.5 kg/m^2^), Normal (BMI 18.5-24.9 kg/m^2^), and Overweight (BMI >24.9 kg/m^2^).(32) A dichotomous variable was then created and labeled as “1” for DBM present (coexistence of maternal overweight and child stunted, underweight, or wasted) and “0” for DBM absent.

#### Independent variables

The selection of independent variables followed the theoretical framework proposed by Hien and Hoa (2009) (**Supplementary Figure S2**), guided by their relevance and theoretical linkage to DBM.(33–35) The framework conceptualizes how DBM can be influenced by socio-demographic, maternal, environmental, and proximal factors. Independent variables considered in this study include- maternal age, education, marital status, employment status, number of children born alive. We also evaluated some child-related variables- child’s age, sex, birth weight, history of recent illness (two weeks prior to data collection), such as fever, cough, diarrhoea. Details of these variables and their categorization are presented in the **supplementary table S2**.

#### Women’s empowerment

To generate women’s empowerment index, we have used a modified version of recently developed SWPER Global index.(36) There were overlaps between the original SWPER index and our study variables, so we have decided to use a modified version of this scale. In this modified version we have used a set of 12 variables that capture all three distinct areas of women empowerment: decision making, social independence, and attitude towards violence. Instead of creating three separate women empowerment index scores, we have created one composite score using principal component analysis and further categorized into five-quintiles similar to the household wealth index. Details of these variables and their categorization are presented in the supplementary **table S1**.

#### Household characteristics

Several household related variables such as place of residence (urban or rural), household wealth status, source of drinking water, toilet facilities and region were included in this analysis. The wealth index is a measure of household’s living standard. This variable was calculated using data on a household’s ownership of a number of selected assets such as radio, television, bicycle, and car; electricity; type of materials used for construction of dwelling, such as flooring material; type of stove, water access, and sanitation facilities and other characteristics related to wealth status. Then the index is generated by principal component analysis and the score is grouped into five categories: poorest, poorer, middle, richer, and richest quintiles.(17) The improved drinking- water sources include household connections (piped to dwelling, yard/plot, piped to neighbour), public tap/standpipes, tubewells/boreholes, protected dug wells, protected springs, and rainwater collection. Unimproved water sources include unprotected wells, unprotected springs, surface water (e.g. rivers, ponds, dam or lakes), bottled water, and tanker truck-provided water.(16) Toilet facilities were considered improved if included flush to piped sewer system, septic tank, ventilated-improved pit latrines, or pit latrines with slab. Flush to elsewhere else, pit latrines without slabs or open pits, no flush to piped sewer system or septic tank or open defecation were not considered as improved sanitation.(16)

### Statistical analysis

We followed “Guide to DHS statistics-8” for this analysis.(31) The sample characteristics were presented with frequencies and percentages of the study variables. To identify the factors associated with DBM, bivariate logistic regression analyses were first conducted, and variables with a p-value <0.20 were considered for inclusion in the multivariable model. A multivariable binary logistic regression model was then constructed to estimate adjusted association between explanatory variables and DBM. Results were reported as odds ratios (OR) with 95% confidence intervals (CIs). Model fitness was assessed using the Hosmer–Lemeshow goodness-of-fit test. Pre-specified interaction terms were tested to assess whether the association between women’s empowerment and DBM varied by key maternal characteristics (age, education, and employment status).

The Concentration Index (CI) and Concentration Curve (CC) approaches were followed to identify household wealth and women empowerment-related inequalities in DBM.(15) The value of CI ranges between +1 and -1, with a zero-value indicating no socioeconomic inequalities. A negative value indicates unequal concentration of outcome variable among the poor (pro-rich inequalities), which can be visualized in CC as the segment below the 45-degree reference line and vice-versa (pro-rich inequality; CC above the 45-degree reference line).(37) Due to binary nature of our outcome variable, Erreygers’s correction approach was followed to bound the CI value between +1 to -1(19), which was also followed in previous research.(38)

Data were analyzed using Stata statistical software version 18.0.(39) Sampling weights were applied to ensure representativeness of the sample. Statistical significance was considered at the p<0.05 level.

### Ethical considerations

This study used data from Mozambique Demographic and Health Survey 2022-2023. The survey was approved by National Bioethics Committee for Health (CNBS) and ICF international institutional review board (IRB). Further approval for this study was not required since data is freely available in the public domain.

## RESULTS

### 3.1 Sample characteristics

**Table 1** show characteristics of the mother-child dyads. Majority of mothers (67.3%, n = 2398) were aged 20-34 years. Nearly half of the mothers (48.2%, n = 1696) had completed primary education, the majority were not employed (72.8%, n = 2435) and married (82.7%, n = 2941). Child characteristics showed a relatively even distribution across the age groups from 0-59 months, with a slightly higher proportion in the 13-24 months (21.4%, n = 772). Nearly equal distribution was observed among male (48.4%, n = 1787) and female (51.6%, n = 1818) children. Household characteristics indicated predominantly rural residence (71.2%, n = 2481). Regional distribution varied considerably, with the highest representation from Nampula (27.1%, n = 487) province.

**Table 1.** Distribution of sample characteristics: Mother-child dyads from Mozambique Demographic and Health Survey, 2022-2023.

| Characteristics | Frequency | Weighted proportion (%) |
| --- | --- | --- |
| **Maternal characteristics** |  |  |
| **Maternal age** |  |  |
| 15-19 years | 381 | 9.6 |
| 20-34 years | 2398 | 67.3 |
| 34 and above | 826 | 23.1 |
| **Maternal educational status** |  |  |
| No formal education | 1026 | 31.2 |
| Primary | 1696 | 48.2 |
| Secondary | 817 | 19.4 |
| Higher | 66 | 1.2 |
| **Employment status** |  |  |
| Not working | 2435 | 72.8 |
| Working | 1170 | 27.2 |
| **Marital status** |  |  |
| Never in union | 213 | 5.1 |
| Married/Living with partner | 2941 | 82.7 |
| Other (Widowed/Divorced/Separated) | 451 | 12.2 |
| **Number of children born alive** |  |  |
| 1 | 629 | 16.3 |
| 2-3 | 1380 | 37.7 |
| 4+ | 1596 | 46.0 |
| **Women Empowerment Index** |  |  |
| Lowest (1<sup>st</sup> quintile) | 589 | 23.9 |
| Lower (2<sup>nd</sup> quintile) | 610 | 23.9 |
| Moderate (3<sup>rd</sup> quintile) | 834 | 27.9 |
| High (4<sup>th</sup> quintile) | 598 | 16.8 |
| Highest (5<sup>th</sup> quintile) | 310 | 7.5 |
| **Child characteristics** |  |  |
| **Age of the child** |  |  |
| <6 months | 515 | 14.4 |
| 6-12 months | 414 | 11.0 |
| 13-24 months | 772 | 21.4 |
| 25-36 months | 675 | 19.2 |
| 37-48 months | 651 | 17.4 |
| 49-59 months | 578 | 16.6 |
| **Sex of the child** |  |  |
| Male | 1787 | 48.4 |
| Female | 1818 | 51.6 |
| **Birth weight** |  |  |
| Low birth weight (<2.5 kg) | 188 | 4.7 |
| Normal (2.5-4.5 kg) | 1316 | 33.1 |
| High birth weight (>4.5 kg) | 815 | 26.5 |
| Not applicable | 1286 | 35.7 |
| **History of recent illness** |  |  |
| **Fever** |  |  |
| No | 3150 | 89.1 |
| Yes | 455 | 10.9 |
| **Cough** |  |  |
| No | 3133 | 90.5 |
| Yes | 472 | 9.5 |
| **Diarrhoea** |  |  |
| No | 3215 | 91.0 |
| Yes | 390 | 9.0 |
| **Household characteristics** |  |  |
| **Place of residence** |  |  |
| Urban | 1124 | 28.8 |
| Rural | 2481 | 71.2 |
| **Household wealth status** |  |  |
| Poorest (1<sup>st</sup> quintile) | 784 | 26.8 |
| Poorer (2<sup>nd</sup> quintile) | 670 | 22.2 |
| Middle (3<sup>rd</sup> quintile) | 815 | 19.1 |
| Richer (4<sup>th</sup> quintile) | 743 | 19.0 |
| Richest (5<sup>th</sup> quintile) | 593 | 12.9 |
| **Source of drinking water** |  |  |
| Improved | 2241 | 56.3 |
| Not improved | 1323 | 43.7 |
| **Type of toilet** |  |  |
| Improved | 920 | 26.0 |
| Not improved | 2644 | 74.0 |
| **Region/Provinces** |  |  |
| Niassa | 428 | 8.7 |
| Cabo Delgado | 479 | 6.8 |
| Nampula | 487 | 27.1 |
| Zambezia | 279 | 17.5 |
| Tete | 342 | 10.4 |
| Manica | 381 | 7.6 |
| Sofala | 361 | 7.0 |
| Inhambane | 229 | 3.4 |
| Gaza | 258 | 3.7 |
| Maputo | 203 | 5.7 |
| Cidade de Maputo | 158 | 2.1 |

### 3.2 Prevalence of dual burden of malnutrition in mother-child dyads in Mozambique

**Table 2** shows the pattern of malnutrition among mothers and children in Mozambique. Maternal malnutrition indicators showed that while most mothers (74.3%) had normal BMI, 6.2% were underweight and 19.5% were overweight/obese. Among children, more than one-third of the children (37.3%) had at least one form of undernutrition (stunting, underweight, or wasting). The combined mother-child nutritional profile demonstrated that 48.7% of the dyads had no malnutrition, while 31.8% of the dyads exhibited child undernutrition without maternal overweight (BMI: normal/underweight). Maternal overweight without child undernutrition was present in 14.0% of the dyads. Notably, 5.5% of the pairs demonstrated DBM, characterized by concurrent maternal overweight/obese condition and child undernutrition (either stunting, underweight, or wasting) in the same household. **Figure 1** illustrates the geographical distribution of DBM among mother-child dyads in Mozambique.

**Figure 1:**
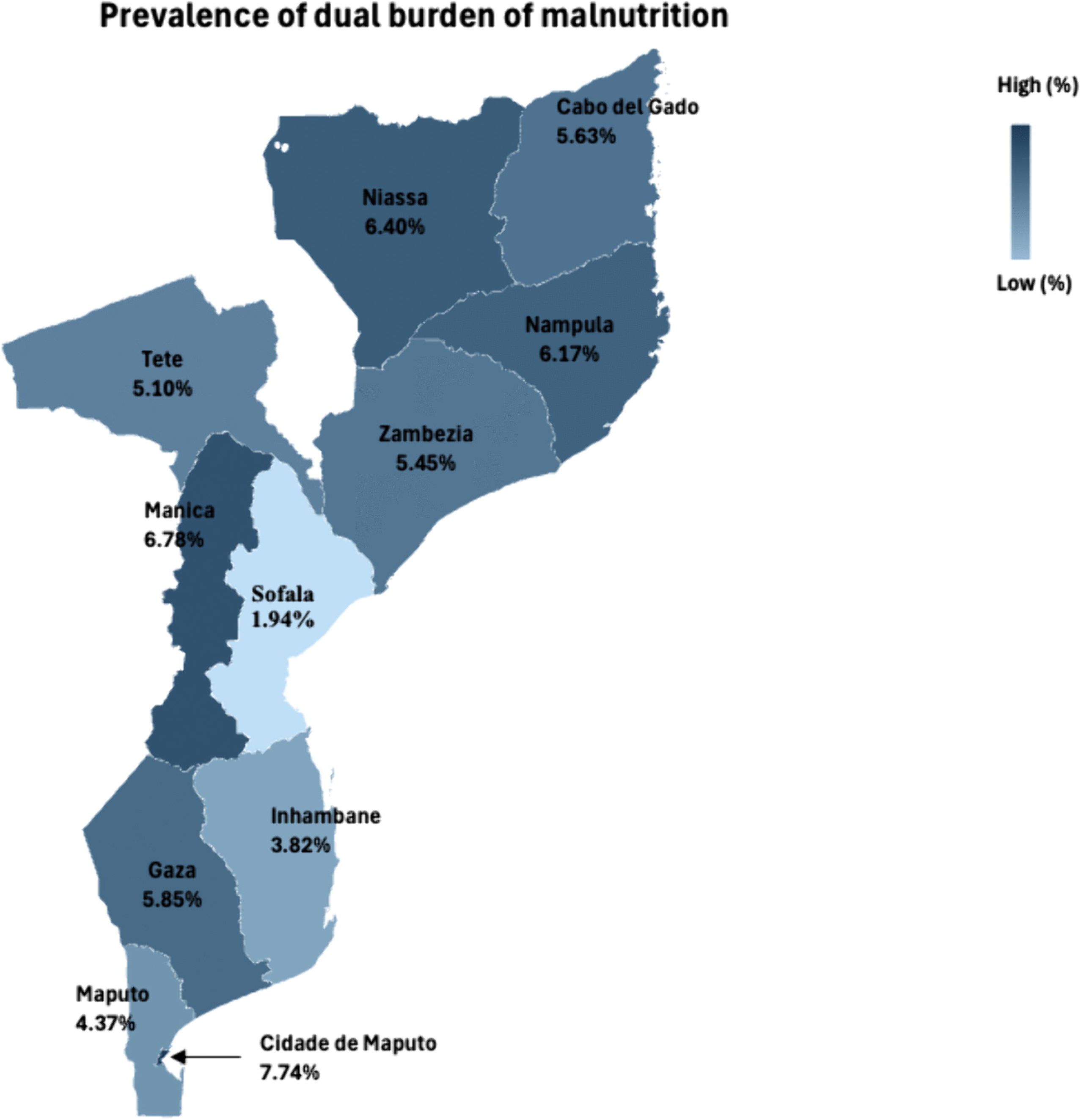
**Geographical distribution of dual burden of malnutrition among mother-child dyads in Mozambique**

**Table 2:** Prevalence of nutritional burden among mothers and children in Mozambique: DHS 2022-2023.

| Characteristics | Frequency | Weighted proportion (%) |
| --- | --- | --- |
| **Maternal BMI status** |  |  |
| Underweight | 181 | 6.2 |
| Normal | 2632 | 74.3 |
| Overweight/Obese | 792 | 19.5 |
| **Undernutrition condition of children** |  |  |
| No | 2362 | 62.7 |
| Yes (either stunting/wasting/underweight ) | 1243 | 37.3 |
| **Combined Nutritional status of mother-child dyads** |  |  |
| Normal nutritional status | 1762 | 48.7 |
| Maternal overweight, no child undernutrition | 600 | 14.0 |
| Child undernourished, no maternal overweight | 1051 | 31.8 |
| Maternal overweight and child undernutrition present | 192 | 5.5 |
| **Dual burden of malnutrition** |  |  |
| No | 3413 | 94.5 |
| Yes | 192 | 5.5 |

### 3.3 Factors associated with the dual burden of malnutrition among mother-child dyads in Mozambique

In bivariate analysis, the Chi-square test revealed a significant association between DBM and several independent variables such as maternal age, education, number of children ever born, women empowerment index, age of child, sex of child, birth weight, place of residence, household wealth status, source of drinking water, and type of toilet (**Supplementary Table S3**). Findings from the most parsimonious multivariable logistic regression model suggest that maternal age, number of children, age of the child, place of residence, and type of toilet facilities were significantly associated with DBM (**Table 3**). Mothers aged >34 years were 4 times more likely to have DBM [95% CI: 1.44, 11.14; p<0.05] compared to those aged 20-34 years. The likelihood of DBM increased with the number of children born. Mothers with 4 or more children had significantly higher odds of experiencing DBM, with an odds ratio of 2.68 [95% CI: 1.29, 5.57; p < 0.05], compared to mothers with one child. Furthermore, children aged 25-36 months [aOR (95% CI): 1.95 (1.00, 3.79); p <0.05] and children aged 37-48 months [aOR (95% CI): 2.17(1.03,4.61); p<0.05] were more likely to be classified as DBM compared to children <6 months.

**Table 3:** Results from final multivariable binary logistic regression model for factors associated with dual burden of malnutrition among mother-child dyads in Mozambique: Demographic and Health Survey, 2022-2023.

| Characteristics | Dual burden of malnutrition |  |
| --- | --- | --- |
|  | aOR<br>[95% CI] | p-value |
| **Maternal characteristics** |  |  |
| **Maternal age** |  |  |
| 20-34 years | Ref |  |
| 15-19 years | 7.33<br>[0.34, 156.16] | 0.20 |
| 34 and above | 4.01<br>[1.44, 11.14] | **<0.05** |
| **Number of children born alive** |  |  |
| 1 | Ref |  |
| 2-3 | 1.94<br>[0.97, 3.87] | 0.06 |
| 4 or more | 2.68<br>[1.29, 5.57] | **<0.05** |
| **Women Empowerment Index** |  |  |
| Highest (5<sup>th</sup> quintile) | Ref |  |
| Lowest (1<sup>st</sup> quintile) | 1.70<br>[0.69, 4.20] | 0.25 |
| Lower (2<sup>nd</sup> quintile) | 0.96<br>[0.39, 2.41] | 0.94 |
| Moderate (3<sup>rd</sup> quintile) | 1.62<br>[0.66, 3.97] | 0.29 |
| High (4<sup>th</sup> quintile) | 1.64<br>[0.71, 3.78] | 0.25 |
| **Age of the child** |  |  |
| <6 months | Ref |  |
| 6-12 months | 0.57<br>[0.20, 1.63] | 0.29 |
| 13-24 months | 1.76<br>[0.88, 3.46] | 0.12 |
| 25-36 months | 1.95<br>[1.00, 3.79] | **<0.05** |
| 37-48 months | 2.17<br>[1.03, 4.61] | **<0.05** |
| 49-59 months | 1.61<br>[0.76, 3.39] | 0.21 |
| **Place of residence** |  |  |
| Urban | Ref |  |
| Rural | 0.51<br>[0.33, 0.79] | **<0.05** |
| **Type of toilet (n=3564)** |  |  |
| Improved | Ref |  |
| Not improved | 0.60<br>[0.38, 0.91] | **<0.05** |

Note: Significant p-value <0.05 are marked in bold.

According to the location of residence, DBM was less likely to be experienced by the mother-child dyads from rural areas [aOR (95% CI): 0.51 (0.33, 0.79); p <0.05] compared to urban residents. Respondents with unimproved toilet facilities were less likely to have DBM [aOR (95% CI): 0.60 (0.38-0.91); p <0.05] compared to those with improved toilet facilities.

#### Hosmer-Lemeshow Goodness-of-fit test significance at p= 0.93 indicated good fit of the model

A significant effect modification was observed between women’s empowerment and maternal age (p <0.05), indicating that the probability of experiencing DBM varies across different age groups and empowerment levels. Younger mothers, aged 15-19 years, with either lowest or highest empowerment levels were more likely to experience DBM compared to mothers aged 20-34 years with highest empowerment level (**Figure 2**).

**Figure 2:**
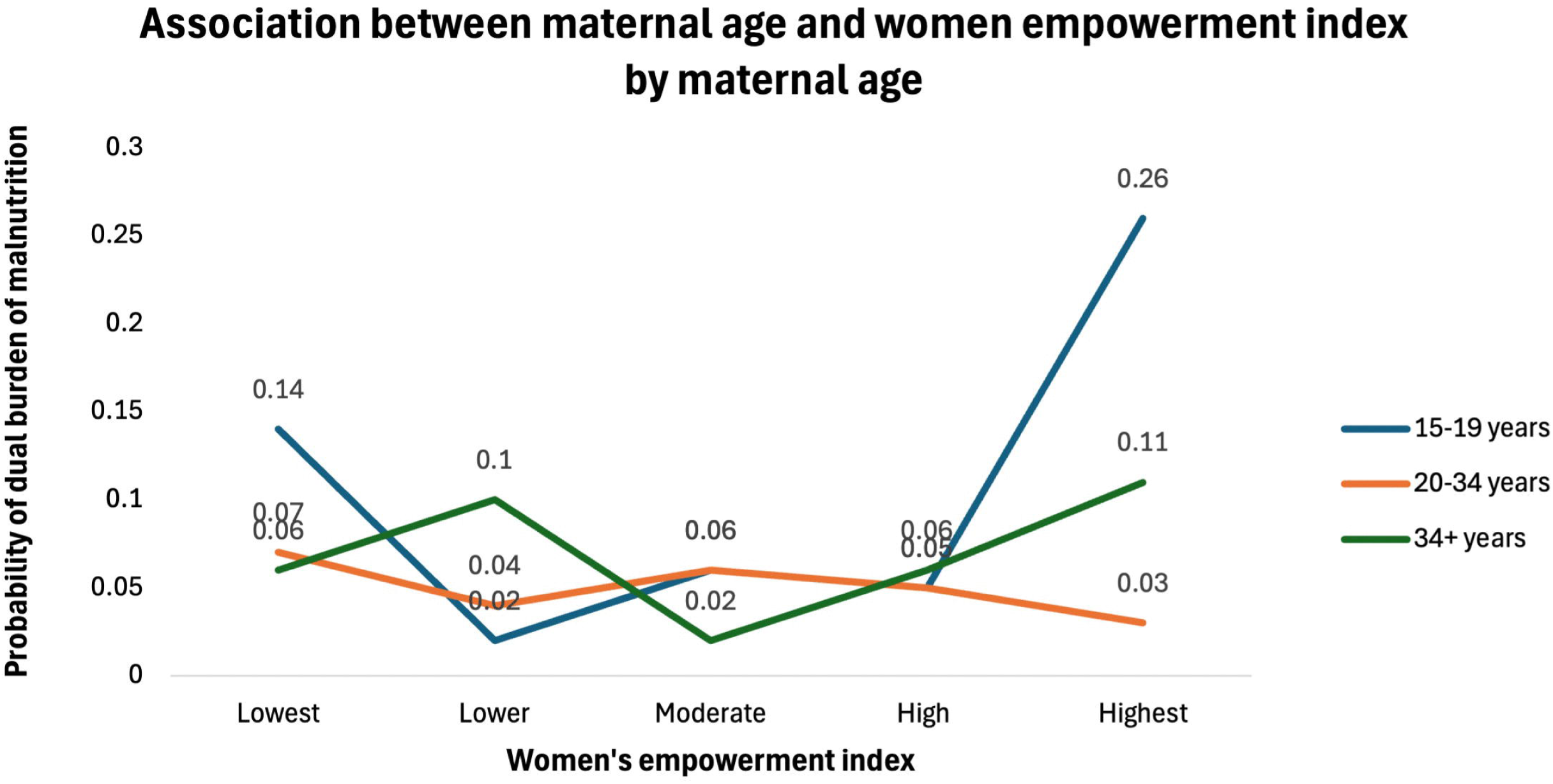
**Effect modification plot showing association between women’s empowerment and DBM modified by maternal age groups**

### 3.4 Inequalities in dual burden of malnutrition

The household wealth and women’s empowerment-related inequalities in DBM among mothers and children in Mozambique are presented in **supplementary table S4**. A significant disparity was observed by Erreygers’s concentration index (ECI) for household wealth (ECI (SE): 0.05(0.01); p<0.001). The positive value of ECI indicates that the inequality in DBM was observed among higher wealth quintiles. The Concentration Curves (CC), showing that the estimated curve lies below the line of equity, indicating the presence of wealth related inequalities in DBM. However, ECI for women’s empowerment index was not significant for DBM (**Supplementary Figure S3**).

## DISCUSSION

This study has systematically identified the breadth of sociodemographic factors and social inequalities associated with dual burden of mother-child overweight and malnutrition in the same household, using nationally representative data from Mozambique. We found that maternal age, number of children born alive, age of the child, place of residence, and type of toilet facilities, reported a positive association with DBM.

In our study, the prevalence of DBM was approximately 5.5% amongst mother-child dyads. This finding is consistent with evidence from sub-Saharan Africa showing that maternal overweight and child undernutrition increasingly co-occur within households, reflecting the region’s ongoing nutrition transition.(40) The issue of DBM amongst mother-child dyads is not an isolated phenomenon.(41) Rather, it is influenced by the rapid increase in maternal weight and a slower reduction rate in child malnutrition. These findings are in line with previous studies conducted in Asia and sub-Saharan Africa where the coexistence of undernutrition and overnutrition has been observed.(26,42–44)

An interesting finding from this study is that whilst most mothers (74.2%) were deemed to have a normal BMI (higher than that of countries like Sierra Leone (62.2%)), 6.2% were underweight, and 19.5% were overweight/obese.(45) These results warrant cautious interpretation. BMI calculation, which squares height to minimize leg length’s influence, assumes mass concentration in the trunk. Its utility is limited for individuals of short stature and does not account for body type variations between individuals.(46) Zierle-Ghosh and Jan (2023) (46) noted BMI’s effectiveness in identifying obesity, but its limitations in ruling it out.(46,47) Such nuances bear greater clinical than policy relevance. Our study, however, did not directly use anthropometric measures to determine individuals’ nutritional status. Despite its limitations, BMI remains the most widely used tool for evaluating individual nutritional status.(47–49)

Overall, factors such as lifestyle choices, living conditions, poverty, limited access to resources and services, diseases, inadequate and inconsistent income populated low-communities and aging may play a role.(50,51) Onyango and colleagues (2019) (40) suggested that the drivers of DBM originate from outside the health sector and operate across national and regional boundaries. For example, in South Africa, which borders Mozambique in the south, being overweight is not perceived in the same way amongst different ethnic groups and across the border. In many African communities, a larger waist symbolizes beauty, prosperity, well-being and good health. As a result, this perception amongst women may explain the societal context of higher prevalence of overweight and obesity.(52) Onyango et al. (2019)(40) further argued that poverty, hunger, and disease are central contributors to malnutrition in Africa, closely linked to insecure livelihoods, poor living conditions, limited education, and insufficient access to essential healthcare services and nutritious food.

Regarding age of the mother, the study revealed that older mothers (>34 years old) were significantly more likely to experience DBM (4.01; 95% CI 1.44 to 11.14), compared to younger mothers (20-24 years). These findings are consistent with that of existing literature, which reported that advanced maternal age was associated with a higher likelihood of experiencing DBM compared to younger age groups.(50,53) Furthermore, as age increases, the risk of diseases, e.g., dietary-related NCDs rises significantly, heightening the potential for nutrition-related health problems. Our findings are also consistent with those of Sunuwar et al. (2020), who reported that mothers above 35 years-old were found to be at higher risk of experiencing DBM.(42)

In our study, over one-third of the children were affected by some form of undernutrition (stunting, underweight, or wasting). Children aged 25-36 months and 37-48 months (1.92; 95% CI 0.99 to 3.68); (2.08; 95% CI 0.99 to 4.39) had higher odds of experiencing DBM than children younger than 6 months, respectively. DBM can be observed at multiple levels, including both population and individual levels.(54) It is also observable within communities and households, particularly where children deemed stunted live in households with overweight mothers.(53) A study conducted in the East and Southern African regions found that the overall stunting prevalence was 30.5%, with prevalence ranging from 22.3% in Namibia to 42.8% in Mozambique. It also revealed that 23% (3 out of 13) of the East and Southern African countries including Comoros, Rwanda, and Mozambique had prevalences of overnutrition as well as undernutrition, exceeding the regional average prevalence estimates for both conditions.(54)

Further analysis also showed that the place of residence exacerbates DBM amongst mother-child dyads. Compared to mothers residing in urban areas, those in rural areas typically had a lower DBM. A variety of factors could potentially cause this phenomenon. First and foremost, rural residents are often engaged in physically demanding activities like farming, food preparation such as grinding/pounding grains, and selling produce on the streets or local markets. This may lead to higher prevalence of rural mothers engaged more in physical activity.(55,56) Consequently, this might help prevent weight gain and lower the prevalence of metabolic disorders amongst women living in rural areas. Conversely, possible reasons for the association between urban dwellers and overweight or obesity may be that urban dwellers are more likely to consume ready-made, packaged and deep-refrigerated foods and adopt a sedentary lifestyle.(50) Our findings align with existing research conducted in Tanzania, China, Bangladesh, and in the sub-Sahara African region, which showed that the prevalence of overweight and obesity was significantly higher in urban areas compared to rural areas.(2,57,58) As a result, urban families are more susceptible to metabolic disorders due to an increased incidence of obesity. Nevertheless, this circumstance does not apply equally to all urban children as they generally have a lower likelihood of experiencing any form of undernutrition compared to their rural counterparts.(55)

In our study, maternal age modified the association between women’s empowerment and DBM of mother-child dyads. Specifically, among younger mothers aged 15-19 years, those at either the lowest or highest levels of empowerment were more likely to experience DBM compared to mothers aged 20-34 years old with the highest empowerment level. This finding is surprising-especially higher risk of mother-child DMB among young mothers who are deemed to express highest level of empowerment-and we question whether, in this instance, this is due to an artefact of measurement; that is, measurement of women’s empowerment may have been mis-applied to women in the youngest of age groups (15-19 years). Teen-aged mothers with children, while not uncommon, raise their child(ren) in socially, economically, and developmentally complex and challenging circumstances. Questions that comprise women’s empowerment index may be differentially suited to the teen mothers with child compared to that for older mothers.

A prior study revealed that mother-child dyads in the wealthiest households have a higher risk of DBM compared to those in the poorest households.(55) One possible cause of these disparities is mothers’ dietary practices. Mothers from affluent households may be more inclined to choose meals insufficient in essential micronutrients yet high in fat, and refined sugars, thereby increase their susceptibility to overweight or obesity.(55) In our study, mothers in wealthy households had higher risk for DBM (1.36; 95% CI 0.51 to 3.65). This finding is corroborate with previous studies as well.(49,50,55) In our study, a significant disparity was observed for household wealth. The positive concentration index indicates that inequality in DBM was observed amongst higher wealth quintiles. Households classed as middle, richer, and richest have significantly higher odds of experiencing DBM compared to the poorest households. These findings suggest that middle- and high-income households in Mozambique may be at a greater risk for DBM, which aligns with findings from previous studies.(41,57,59)

Mozambique’s rapid economic expansion before 2015, largely fueled by extractive industries and megaprojects, positioned the country among the fastest-growing economies globally. However, the benefits of this growth were unequally distributed, primarily accruing to the wealthiest 20% of the population, while poverty deepened among the poorest majority. Despite sustained, though slower, growth after 2015, extreme poverty increased, with nearly three-quarters of the population living on less than $2.15 a day by 2019.(60) This economic paradox--high growth with rising poverty and inequality-has important implications for health and nutrition. As incomes and urbanization rise among a minority, dietary transitions toward energy-dense, processed foods are observed, while widespread poverty sustains undernutrition. This coexistence of ‘overnutrition’ in wealthier groups and persistent undernutrition in vulnerable populations underpins DBM in Mozambique, reflecting an ongoing epidemiological and nutritional transition shaped by uneven economic development. Like many other LMICs, Mozambique has made remarkable progress in curtailing undernutrition rates in the past decade. (40,54) At the same time, rising rates of overweight and obesity, linked to the country’s ongoing epidemiological and nutritional transition, are placing new pressures on the National Health System. Communities are caught in a dual burden, where infectious diseases and nutrient deficiencies persist alongside increasing diet-related NCDs. This manifest at both population and household levels, with some members undernourished (e.g., stunted children or anemic adults) while others are overweight or obese.(61)

Recent studies underscore a swift change in the nutritional health status of adults and pregnant women and evolving dietary preferences.(62,63) These studies provide evidence of an ongoing nutrition transition in Mozambique. The shift towards a greater preference for ultra-processed and energy-dense foods and beverages may lead to a diminished intake of essential nutrients and ultimately contribute to child undernourishment. This dietary shift over the past decades underlies the rising prevalence of DBM in Mozambique. Historically, the most prevalent forms of malnutrition among children under five and adolescents worldwide have been undernutrition and micronutrient deficiencies, including stunting, wasting, underweight, and deficiencies of essential micronutrients. Global trends indicate a rapid rise in childhood overweight and obesity, even in countries still grappling with undernutrition conditions. In reality, overweight and obesity are the only forms of malnutrition that are rising among children under five and adolescents at a global level.(64,65) However, in countries like Mozambique and other LMICs, the shifting in dietary changes is not happening in a similar manner as it is in high-income countries.

Our findings are valid for Mozambique. However, they can be applied to other LMICs where mother-child dyads experience similar outcomes. The issues we identified will need appropriate interventions to address the detrimental effects of malnutrition in Mozambique. Tackling these issues requires implementing actions that combat all forms of malnutrition, including driven agricultural and food policy research and providing evidence-based policy support to the local government and agencies. One way of doing so is to adopt the concept of “double-duty actions,” first introduced in the 2015 Global Nutrition Report (IFPRI, 2015), and has since been gradually utilized in nutrition research to holistically address this complex public health problem.(66) Double-duty actions encompass interventions, programs, policies, and regulations that simultaneously reduce the risk of undernutrition and overweight, obesity, or diet-related NCDs.(5) These actions are not necessarily new initiatives. Rather, adequate measures are already in place to address obesity or undernutrition. However, these measures should be further enhanced to specifically support households affected by the problem of DBM nationwide.

### Strength and limitations

This study has several limitations. The cross-sectional nature of the data analyzed cannot establish causality. The sub-analysis of child underweight, stunting, and wasting could not be done separately due to lower prevalence of these conditions. Due to unavailability of the data, various potential confounders (such as physical activity, cultural influences on caregiving practices and foods, more detailed components of nutritional status like body composition or biochemical or metabolic status) that might affect DBM cannot be included in the analysis. Therefore, further exploration is warranted to ascertain the contribution of these potential determinants to the development of various forms of DBM in Mozambique. Despite such limitations, the strength of the present study was that the data were extracted from a nationally representative large, randomized sample. This study provides evidence on presence of DBM among mother-child dyads in the same households in Mozambique which was not previously reported at national scale. This study also highlights the wealth-related inequalities in the prevalence of DBM in Mozambique. Therefore, the findings from this study provides crucial evidence to develop and implement policies that improve maternal and child health and nutrition in Mozambique.

## CONCLUSION

Our study identified a range of risk factors associated with the nutritional status of women of reproductive age (15-49 years) and children (0-59 months) at the population-level, demonstrating that Mozambique continues to experience DBM. This study highlights significant gaps in understanding the various manifestations of this form of malnutrition, particularly concerning mother-child dyads. We found a very high prevalence of DBM when considering social inequalities and factors related to under- and overnutrition amongst mothers and their children in Mozambique. Our findings show that DBM remains a pressing public health problem that needs immediate attention. The study findings will assist policymakers in identifying individuals at risk of DBM and enabling the development and implementation of targeted programs. Mozambique has a robust policy framework to address malnutrition in all its forms. By incorporating the double-duty actions in the country’s programmatic strategies, the country can make steady progress toward achieving the SDGs, Target 2.0.

## Data Availability

The Mozambique Demographic and Health Survey data is available from the DHS data repository which is publicly available.

https://dhsprogram.com/data/available-datasets.cfm

## Acknowledgement

We greatly acknowledge the Demographic and Health Survey (DHS) program for granting access to the Mozambique DHS datasets.

## Author contributions

NM and NS conceptualized this study. NS performed formal analysis of the data. NS (methodology, results) and FA (background, discussion, conclusion) wrote the first draft. All authors reviewed, edited the drafts and approved the final version of the manuscript. NM supervised the work and holds provenance for the article.

## Patient and public involvement

Patients and/or the public were not involved in the design, or conduct, or reporting, or dissemination plans of our study.

## Supporting Information List

**Figure S1.** Selection of sample for the study of dual burden of malnutrition in Mozambique; Demographic and Health Survey 2022-2023

**Figure S2:** Conceptual framework of the determinants of dual burden of malnutrition among mother-child dyads in Mozambique (Source: modified from Hien & Hao,2009 and UNICEF framework)

**Table S1:** Variables considered to generate women empowerment index: Mozambique Demographic and Health Survey Data 2022-2023

**Table S2:** List of Independent variables with categorization

**Table S3.** Findings from bivariate analysis

**Table S4:** Inequalities related to household wealth and women’s empowerment with dual burden of malnutrition among mother-child dyads in Mozambique

**Figure S3:** Concentration curves of dual burden of malnutrition based on household wealth status (a) and women empowerment index (b). Note: C(p) refers to cumulative proportion

