## Supplementary file for "Dual Burden of Malnutrition Among Mother-Child Dyads in Mozambique: Findings from the Demographic and Health Survey 2022-2023"

These supplementary materials are presented in support of the paper:

**Figure S1. Selection of sample for the study of dual burden of malnutrition in Mozambique; Demographic and Health Survey 2022-2023**

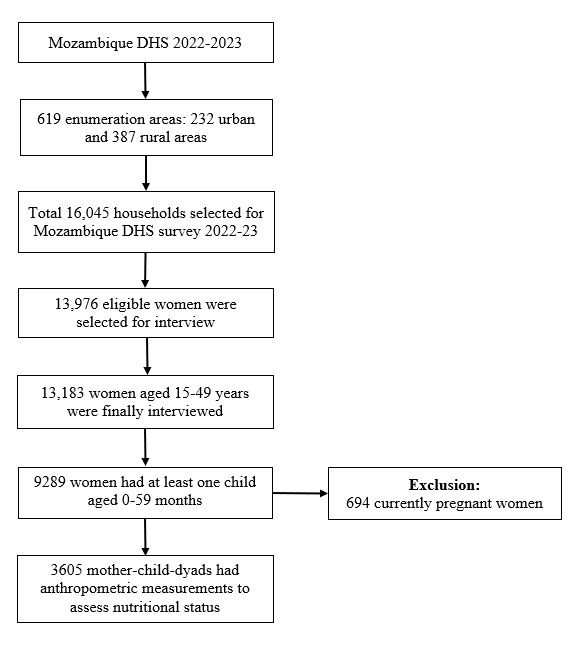

**Figure S2: Conceptual framework of the determinants of dual burden of malnutrition among mother-child dyads in Mozambique (Source: modified from Hien & Hao,2009 and UNICEF framework)**

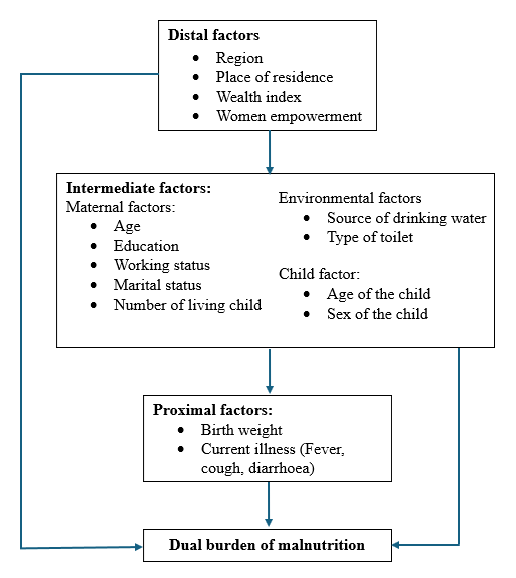

**Table S1: Variables considered to generate women empowerment index: Mozambique Demographic and Health Survey Data 2022-2023**

| **Variable** | **Categorization** | Re-**categorization** |
| --- | --- | --- |
| **Decision making** | | |
| 1. Person who usually decides on: respondent's health care | 1. Alone  2. Jointly with husband/partner  3. Jointly with other person  4. Husband alone  5. Someone else  6. other | - Alone and jointly with husband/partner= 1 - Jointly with other person, husband alone, someone else, other= 0 |
| 2. Person who usually decides on: large household purchases |  |  |
| 3. Person who usually decides on: visits to family or relatives |  |  |
| **Attitude towards violence** | | |
| 4. Beating justified if wife goes out without telling husband | 0. No  1. Yes  8. Don’t know | - No= 1 - Yes and don’t know= 0 |
| 5. Beating justified if wife neglects the children |  |  |
| 6. Beating justified if wife argues with husband |  |  |
| 7. Beating justified if wife refuses to have sex with husband |  |  |
| 8. Beating justified if wife burns the food |  |  |
| **Social independence** | | |
| 9. Respondent reading newspaper/magazine once a week | 0. No  1. Yes | - No= 0 - Yes= 1 |
| 10. Has an account in a bank or other financial institution |  |  |
| 11. Owns a mobile/ telephone |  |  |
| 12. Owns a house/land alone or jointly | 0. Does not own  1. Alone only  2. Jointly with husband/partner  3. Jointly with someone else  4. Jointly with husband/partner and someone else  5. Both alone and jointly | - Does not own= 0 - Own= 1 |

**Table S2: List of Independent variables with categorization**

| **Variable** | **Description** | **Categorization** |
| --- | --- | --- |
| **Maternal characteristics** |  |  |
| Maternal age | Age of the respondents(women) in years | 0. 15-19 years  1. 20-34 years (ref)  2. 35+ years |
| Educational status | Highest educational level completed by the respondents | 0. No formal education (ref) 1. Primary 2. Secondary 3. Higher |
| Employment status | Current working status of the respondents | 0. Not working (ref) 1. Working |
| Marital status | Current marital status of the respondents | 0. Never in union (ref) 1. Married/ Living with partner 2. Other (Widowed/ Divorced/ Separated) |
| Number of children born alive | The number of children the respondent has given birth to. | 0. 0-1 (ref) 1. 2-3 2. 4 or more |
| Women empowerment index | Women empowerment index was developed using principal component analysis. Details about this index are provided in Supplement table S1 | 1. Lowest (Q1) 2. Lower(Q2) 3. Moderate(Q3) 4. High (Q4) 5. Highest (Q5) (ref) |
| **Child characteristics** |  |  |
| Age of the child | Age of the child in months | 0. <6 months (ref) 1. 6-12 months 2. 13-24 months 3. 25-36 months 4. 37-48 months 5. 49-50 months |
| Sex of the child | Sex (at birth) of the child | 0. Male (ref) 1. Female |
| Birth weight | Birth weight of the child in kilograms | 0. Low birth weight (<2.5 kg) 1. Normal (2.5-4.5 kg) (ref) 2. High birth weight (>4.5 kg) |
| **History of recent illness** |  |  |
| Fever | Whether the child had Fever in last 2 weeks of interview | 0. No (ref) 1. Yes |
| Cough | Whether the child had suffered from a cough in last 2 weeks of interview | 0. No (ref) 1. Yes |
| Diarrhoea | Whether the child had diarrhoea in last 2 weeks of interview | 0. No (ref) 1. Yes |
| **Household characteristics** |  |  |
| Place of residence | Type of place of residence | 0. Urban (ref) 1. Rural |
| Wealth status | Household wealth status | 0. Poorest (ref) 1. Poor 2. Middle 3. Richer 4. Richest |
| Source of drinking water | The main source of drinking water for members of the household | 0. Improved (ref) 1. Nor improved |
| Type of toilet | The type of toilet facility the members of the household usually use | 0. Improved (ref) 1. Not improved |
| Region | Administrative regions | 0. Niassa 1. Cabo Delgado 2. Nampula 3. Zambézia 4. Tete 5. Manica 6. Sofala 7. Inhambane 8. Gaza 9. Maputo (ref) 10. Cidade de Maputo |

**Table S3. Findings from bivariate analysis**

| **Characteristics** | **Dual burden of malnutrition** | |
| --- | --- | --- |
|  | **Present**  **% [95% CI]** | **p-value** |
| **Maternal characteristics** | | |
| **Maternal age** |  | 0.145 |
| 15-19 years | 3.42  [1.67, 6.90] |  |
| 20-34 years | 5.30  [4.22, 6.63] |  |
| 34 and above | 6.98  [5.02, 9.64] |  |
| **Maternal educational status** |  | 0.065 |
| No formal education | 4.86  [3.39, 6.93] |  |
| Primary | 5.10  [3.92, 6.62] |  |
| Secondary | 6.97  [4.97, 9.70] |  |
| Higher | 14.47  [6.24, 30.06] |  |
| **Employment status** |  | 0.583 |
| Not working | 5.35  [4.28, 6.66] |  |
| Working | 5.94  [4.39, 7.98] |  |
| **Marital status** |  | 0.352 |
| Never in union | 5.69  [2.44, 12.72] |  |
| Married/Living with partner | 5.76  [4.74, 6.98] |  |
| Other(Widowed/Divorced/Seperated) | 3.73  [2.30, 5.97] |  |
| **Number of children born alive** |  | 0.161 |
| 1 | 4.04  [2.46, 6.57] |  |
| 2-3 | 5.01  [3.72, 6.72] |  |
| 4+ | 6.43  [5.04, 8.18] |  |
| **Women Empowerment Index** |  | 0.064 |
| Lowest (Q1) | 5.90  [3.78, 9.11] |  |
| Lower (Q2) | 4.64  [2.99, 7.15] |  |
| Moderate (Q3) | 4.30  [2.88, 6.37] |  |
| High (Q4) | 8.14  [5.65, 11.59] |  |
| Highest (Q5) | 8.95  [5.65, 13.90] |  |
| **Child characteristics** | | |
| **Age of the child** |  | **0.006** |
| <6 months | 3.26  [1.95, 5.42] |  |
| 6-12 months | 2.04  [0.95, 4.35] |  |
| 13-24 months | 5.26  [3.66, 7.52] |  |
| 25-36 months | 7.50  [5.37, 10.39] |  |
| 37-48 months | 7.55  [5.30, 10.64] |  |
| 49-59 months | 5.60  [3.47, 8.92] |  |
| **Sex of the child** |  | 0.074 |
| Male | 6.43  [5.06, 8.14] |  |
| Female | 4.64  [3.53, 6.07] |  |
| **Birth weight** |  | **0.0342** |
| <2.5 kg (Low birth weight) | 7.78  [4.62, 12.82] |  |
| 2.5-4.5 kg (Normal) | 5.34  [4.03, 7.06] |  |
| >4.5 kg (High birth weight) | 3.75  [2.54, 5.50] |  |
| Not applicable | 6.66  [5.12, 8.63] |  |
| **History of recent illness** |  |  |
| **Fever** |  | 0.274 |
| No | 5.33  [4.41, 6.43] |  |
| Yes | 6.95  [4.40, 10.81] |  |
| **Cough** |  | 0.134 |
| No | 5.30  [4.36, 6.42] |  |
| Yes | 7.52  [4.90, 11.36] |  |
| **Diarrhoea** |  | 0.968 |
| No | 5.51  [4.59, 6.61] |  |
| Yes | 5.44  [2.90, 9.99] |  |
| **Household characteristics** | | |
| **Place of residence** |  | **<0.001** |
| Urban | 8.67  [6.64, 11.25] |  |
| Rural | 4.23  [3.30, 5.40] |  |
| **Household wealth status** |  | **<0.001** |
| Poorest | 3.25  [1.93, 5.43] |  |
| Poorer | 4.19  [2.69, 6.49] |  |
| Middle | 5.88  [4.07, 8.42] |  |
| Richer | 6.66  [4.63, 9.50] |  |
| Richest | 10.17  [7.41, 13.81] |  |
| **Source of drinking water** |  | **0.013** |
| Improved | 6.56  [5.28, 8.12] |  |
| Not improved | 4.08  [2.96, 5.59] |  |
| **Type of toilet** |  | **0.001** |
| Improved | 8.26  [6.16, 10.98] |  |
| Not improved | 4.50  [3.57, 5.65] |  |
| **Region** |  | 0.534 |
| Niassa | 6.40  [4.18, 9.67] |  |
| Cabo Delgado | 5.63  [3.02, 10.27] |  |
| Nampula | 6.17  [4.20, 8.98] |  |
| Zambézia | 5.45  [3.16, 9.26] |  |
| Tete | 5.10  [2.99, 8.58] |  |
| Manica | 6.78  [4.72, 9.64] |  |
| Sofala | 1.94  [0.87, 4.26] |  |
| Inhambane | 3.82  [2.03, 7.06] |  |
| Gaza | 5.85  [3.28, 10.21] |  |
| Maputo | 4.37  [1.98, 9.36] |  |
| Cidade de Maputo | 7.74  [4.55, 12.86] |  |

*Boldened p-value indicates statistical significance

**Table S4: Inequalities related to household wealth and women’s empowerment with dual burden of malnutrition among mother-child dyads in Mozambique**

|  | **Measure of wealth related inequality** | | **Measure of women empowerment related inequality** | |
| --- | --- | --- | --- | --- |
|  | Erreygers Concentration Index | *p-value* | Erreygers Concentration Index | *p-value* |
| Dual burden of Malnutrition | 0.05 (0.01) | **<0.001** | 0.02 (0.01) | 0.0629 |

**Figure S3: Concentration curves of dual burden of malnutrition based on household wealth status (a) and women empowerment index (b). Note: C(p) refers to cumulative proportion**

(a)

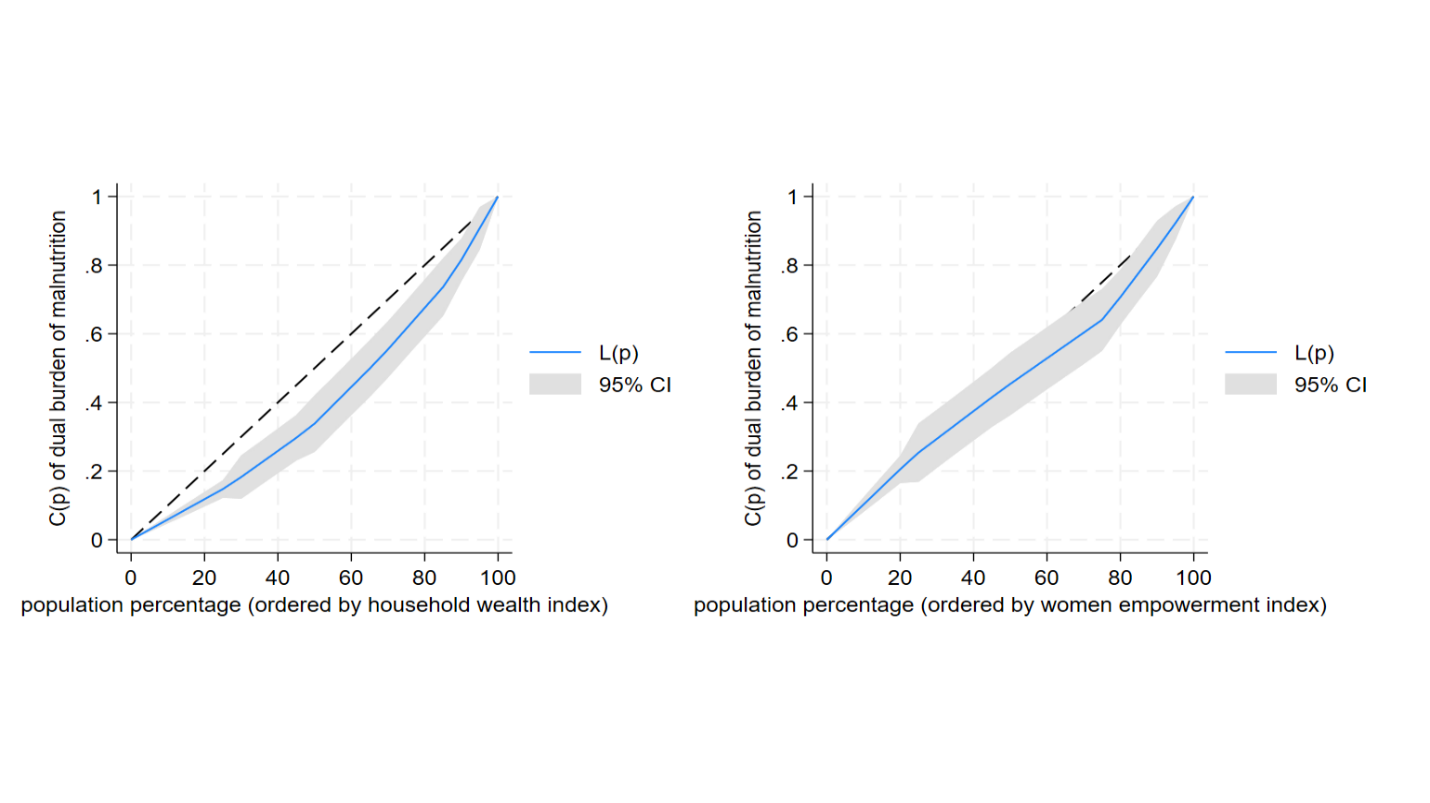

(b)
